# Age disparities in lung cancer survival in New Zealand: the role of patient and clinical factors

**DOI:** 10.1101/2020.10.05.20206912

**Authors:** Sophie Pilleron, Camille Maringe, Hadrien Charvat, June Atkinson, Eva Morris, Diana Sarfati

**Affiliations:** Dept of Public Health, School of medicine, University of Otago, Wellington, New Zealand; Nuffield Department of Population Health, University of Oxford, Big Data Institute, Old Road Campus, Oxford, OX3 7LF, UK; Inequalities in Cancer Outcomes Network, London School of Hygiene and Tropical Medicine, Keppel Street, London WC1E 7HT, UK; Epidemiology and Prevention Group, Center for Public Health Sciences, National Cancer Center, Tokyo, Japan; Section of cancer surveillance, International Agency for Research on Cancer, Lyon, France

**Keywords:** lung cancer, survival, older adults, aged, population-based cancer registry, observational data

## Abstract

**Background:** Age is an important prognostic factor for lung cancer. However, no studies have investigated the age difference in lung cancer survival per se. We, therefore, described the role of patient-related and clinical factors on the age pattern in lung cancer excess mortality hazard by stage at diagnosis in New Zealand.

**Methods:** We extracted 22 487 new lung cancer cases aged 50-99 (median age = 71, 47.1% females) diagnosed between 1 January 2006 and 31 July 2017 from the New Zealand population-based cancer registry and followed up to December 2019. We modelled the effect of age at diagnosis, sex, ethnicity, deprivation, comorbidity, and emergency presentation on the excess mortality hazard by stage at diagnosis, and we derived corresponding lung cancer net survival.

**Results:** The age difference in net survival was particularly marked for localised and regional lung cancers, with a sharp decline in survival from the age of 70. No identified factors influenced age disparities in patients with localised cancer. However, for other stages, females had a greater age difference in survival between middle-aged and older patients with lung cancer than males. Comorbidity and emergency presentation played a minor role. Ethnicity and deprivation did not influence age disparities in lung cancer survival.

**Conclusion:** Sex and stage at diagnosis were the most important factors of age disparities in lung cancer survival in New Zealand.

**Key messages:** *What is the key question?:* How do patient-related and clinical factors influence age pattern in lung cancer survival?

*What is the bottom line?:* Age disparities in lung cancer survival were strongest for females and non-advanced disease. Deprivation, ethnicity, comorbidity, and emergency presentation did not influence age disparities.

*Why read on?:* Our findings reinforce the call for a better representation of older adults in clinical trials and a wider use of geriatric assessment to identify patients who will benefit treatment.

## Introduction

Patients aged 75 years or over have lower lung cancer survival than younger patients^1^. Although lung cancer survival has improved in recent decades, older patients have not benefitted from the improvement in treatment as much as younger patients^1^. The excess mortality in older patients with lung cancer has been observed for up to five years since diagnosis^2^.

Because of the higher likelihood of multiple comorbidities, polypharmacy, age-related physiological changes, frailty, lower functional performance, limited life expectancy^3,4^ and additionally the underrepresentation of older patients in randomised clinical trials, lung cancer management in older patients is challenging5. As a result, older patients with lung cancer have been found to be less likely to receive cancer treatment than younger patients ^6,7^. Also, older adults are more likely to be diagnosed with lung cancer after a presentation to the emergency department, which in turn was associated with advanced cancer and poor survival prospect^8^.

In New Zealand, lung cancer was the third most common cancer diagnosis and the second cause of cancer death in adults aged 70 years and over in 2018^9^. As in other countries, lung cancer five-year survival in patients under the age of 75 has recently improved in New Zealand (from 13.0% (95% confidence interval: 11.9%-14.0%) in 1995-1999 to 18.8% (17.5%-20.2%) in 2010-2014)^1^. However, no improvement was observed for older patients (from 7.9% (6.6%-9.4%) in 1995-1999 to 7.4% (6.2%-8.8%) in 2010-2014), with New Zealanders in this age range having the lowest lung cancer survival among seven high-income countries with similar healthcare system^1^.

Using population-based cancer registry data linked to hospitalisation data, we describe the role of patient-related and clinical factors on the age pattern in lung cancer survival and excess mortality hazard by stage at diagnosis in New Zealand.

## Methods

We included patients aged between 50 and 99 years old at diagnosis, diagnosed with lung cancer (ICD-10 code C34) between 1 January 2006 and 31 July 2017 registered in the New Zealand population-based cancer registry (NZ-PBCR). We restricted our analyses to patients with a first occurrence of lung cancer. We linked these cancer cases to hospitalisation data (National Minimum Dataset) for the five years before the cancer diagnosis, and to the outpatient dataset (National Non-Admitted Patient Collection) where all emergency admissions are recorded. We obtained the date of death for all cancer cases who died between 1 January 2006 and 31 December 2019 from the Ministry of Health.

We categorised the extent of disease as given by NZ-PBCR into Surveillance, Epidemiology and End Results (SEER) stage groups as follows: Localised to the organ of origin or Invasion of adjacent tissue or organ into SEER localised stage; Regional lymph nodes involvement into SEER regional stage; Distant into SEER distant stage; Not known into missing stage^10^.

We used the “all-sites” weighted C3 index developed by Sarfati et al. to assess comorbidity among patients with cancer using administrative hospitalisation data^11^. We defined emergency presentation at diagnosis as a cancer diagnosis occurring in the 28 days following admission to an emergency department^12^.

Sex and age at diagnosis were available for all lung cancer cases within the NZ-PBCR, originating from pathology reports and National Health Index database. We used the New Zealand Deprivation Index (NZDep) to assess the socio-economic deprivation at cancer diagnosis^13^. The NZDep is an ecological indicator built using the nine following variables from the national census: housing tenure, benefit receipt, unemployment, income, telephone access, car access, single-parent families, education and household crowding. The NZdep was assigned to domicile code (census area) of each patient at the time of diagnosis. For all diagnoses occurring during the period 2006-2011, we used the NZDep based on information collected at the 2006 census; for all diagnoses occurring from 2012, we used the score based on information from the 2013 census. We could not map domicile code to an NZDep score for 77 cases; we, therefore, excluded these cases from analyses.

Self-reported ethnicity available in the NZ-PBCR originated from the hospital records. Ethnicity refers to the ethnic group people identify with or feel they belong to, and not to race, ancestry, nationality or citizenship (https://www.stats.govt.nz/topics/ethnicity). Ethnicity data were not available for 86 cases; we, therefore, excluded these cases from analyses.

### Statistical analysis

When studying the survival of cancer patients, it is important to recognize that patients might die from their disease but also from other causes. This becomes crucial when looking at the effect of age at diagnosis as older patients are subjected to a greater risk of dying from competing causes (cardiovascular diseases, etc.). Net survival is the indicator commonly used to analyse population-based cancer registry data when information on cause of death is either missing or deemed unreliable. Net survival is a particularly interesting indicator in this context: it represents the hypothetical survival that would be observed if lung cancer were the only cause of death.

Net survival is derived from the estimation of individual excess mortality hazard (EMH): the overall hazard of death for cancer patients is assumed to be the sum of the expected (or background) hazard and the excess hazard due to cancer. The excess hazard embeds direct (e.g., multiple organ failure resulting from cancer progression) and indirect (e.g., treatment toxicity, pulmonary embolism) effects of cancer on mortality. We used lifetables of mortality in the general population to estimate expected mortality, defined by calendar period (2005-2007 and 2012-2014), single year of age, sex, and ethnicity (Māori, non-Māori) obtained from Statistics New Zealand (http://archive.stats.govt.nz/browse_for_stats/health/life_expectancy/period-life-tables.aspx#gsc.tab=0). We built lifetables for each calendar year between 2006 and 2013 assuming linear interpolation between the age-, sex- and ethnic-specific mortality rates in 2005-7 and 2012-4. We extrapolated the rates beyond 2014 using the same constant yearly change in mortality. We fitted flexible excess hazard regression models for each stage at diagnosis. We censored all patients who were still alive beyond 10 years after diagnosis. The model-building strategy is available in supplemental Material.

All predictions presented in the manuscript were made for Non-Māori males diagnosed in 2012, who lived in an area with third quintile of deprivation index, with median comorbidity score, no emergency admission recorded in the month prior to their cancer diagnosis, unless otherwise stated.

We performed data management using Stata (version 16.0; StataCorp, 2019) and statistical analyses using R statistical software (version 3.4.0; R Development Core Team, 2017), in particular the ‘mexhaz’ package was used for flexible excess hazard modelling^14^.

## Results

Out of 23,645 patients diagnosed with lung cancer in 2006-17 in New Zealand, we included 22,487 (95.1%) patients aged 50-99 years at diagnosis (median age = 71, interquartile range 64-79), 47.1% of whom were females. Table 1 describes patients’ characteristics by age group and supplementary Table 1 by age group and stage at diagnosis. Over half the patients aged 65 years or over were males. Māori represented one-third of lung cancer patients aged 50-64 years, but only 4% in the 85-99 years age group. The number of cases increased with increasing deprivation (supplementary Table 1). Overall, 6.0% of patients were diagnosed with localised cancer, 12.4% with regional cancer, 44.4% with distant cancer, and 37.1% had a missing stage. The percentage of missing stage increased with age at diagnosis. Among patients with known stages, lung cancer was mostly diagnosed at an advanced stage in all age groups. The percentage of patients with at least one comorbidity increased with age from 66% in the 50-64 age group to around 75% after 75 years old. Lung cancer was diagnosed following an emergency presentation in the previous 28 days in 40.6% of patients, and this percentage was highest after the age of 85 (47.7%), and in patients with distant cancer (54.0%).

**Table 1.** Characteristics of patients with lung cancer by age at diagnosis

| Age categories (years) | 50-64 | 65-74 | 75-84 | 85-99 |
| --- | --- | --- | --- | --- |
| Cases (%) | 6062 | 7735 | 6498 | 2192 |
| Male (%) | 2925 (48.3) | 4143 (53.6) | 3632 (55.9) | 1187 (54.2) |
| Ethnicity (%) |  |  |  |  |
| Māori | 1921 (31.7) | 1564 (20.2) | 731 (11.2) | 93 (4.2) |
| Non-Māori | 4141 (68.3) | 6171 (79.8) | 5767 (88.8) | 2099 (95.8) |
| Deprivation index quintiles (%) |  |  |  |  |
| 1- Least deprived | 680 (11.2) | 901 (11.6) | 779 (12.0) | 271 (12.4) |
| 2 | 832 (13.7) | 1071 (13.8) | 952 (14.7) | 396 (18.1) |
| 3 | 1098 (18.1) | 1521 (19.7) | 1360 (20.9) | 499 (22.8) |
| 4 | 1440 (23.8) | 1963 (25.4) | 1758 (27.1) | 565 (25.8) |
| 5 - Most deprived | 2012 (33.2) | 2279 (29.5) | 1649 (25.4) | 461 (21.0) |
| Stage at diagnosis (%) |  |  |  |  |
| Localised | 459 (7.6) | 596 (7.7) | 288 (4.4) | 17 (0.8) |
| Regional | 935 (15.4) | 1089 (14.1) | 656 (10.1) | 109 (5.0) |
| Distant | 3039 (50.1) | 3432 (44.4) | 2649 (40.8) | 874 (39.9) |
| Missing | 1629 (26.9) | 2618 (33.8) | 2905 (44.7) | 1192 (54.4) |
| Comorbidity score >0 (%) | 3977 (65.6) | 5555 (71.8) | 4951 (76.2) | 1631 (74.4) |
| Comorbidity score (median [IQR]) | 0.92 [0.00, 1.84] | 1.09 [0.00, 2.33] | 1.21 [0.11, 2.63] | 1.24 [0.00, 2.76] |
| Emergency route (%) | 2452 (40.4) | 2912 (37.6) | 2718 (41.8) | 1046 (47.7) |

Net survival decreased with more advanced disease (Figure 1): 93.2% (95% confidence interval: 91.7%-94.7%) and 87.8% (85.7%-89.9%) of patients with localised disease survived their cancer beyond 1 and 3 years respectively, 58.1% (56.5%-59.7%) and 33.1% (31.4%-34.8%) of patients with regional cancer, 14.6% (14.0%-15.2%) and 3.9% (3.6%-4.3%) of patients with distant cancer, and 40.4% (39.4%-41.4%) and 14.0% (13.3%-14.7%) of patients with missing stage. Regardless of stage at diagnosis, net survival decreased as age at diagnosis increased (Figure 1 and supplementary Table 2). The age difference in net survival was more marked for localised and regional stages cancers than in other stages, and the smallest difference was observed at three years in patients with distant cancer. We noted a clear decline in survival from the age of 70 for localised and regional disease, but a more gradual decline from the age of 50 for advanced disease.

**Figure 1.**
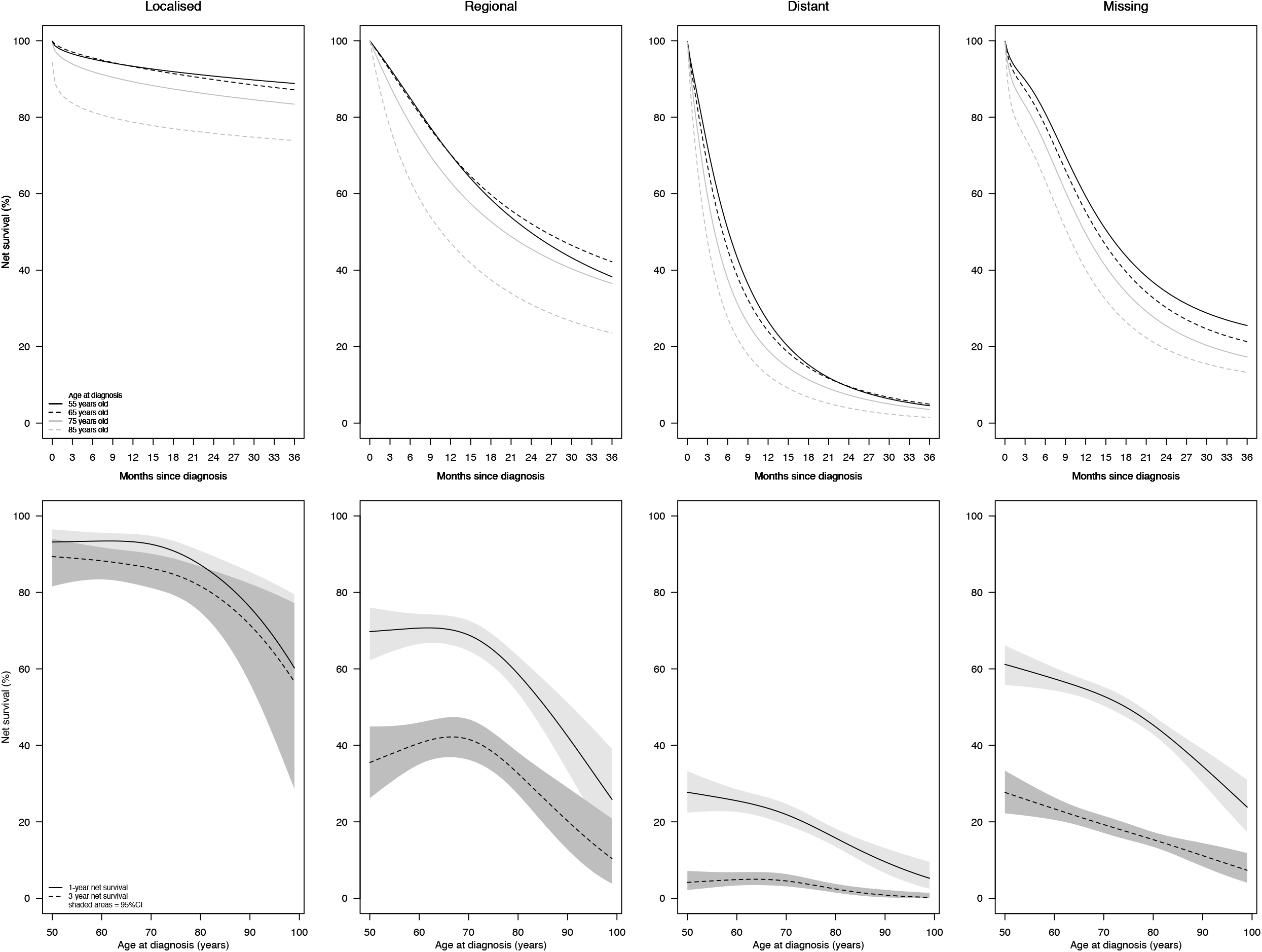
Lung cancer net survival for patients aged 55, 65, 75 and 85 years old over the first three years since diagnosis (top panel), and one-and three-year lung cancer net survival by age at diagnosis (bottom panel) by stage at diagnosis

The age pattern of EMH in patients with lung cancer by stage at diagnosis is presented in Figure 2. The excess risk of death increased with more advanced disease for all ages. In patients with localised cancer, compared with patients aged 55 years old, those aged over 75 years old had higher excess mortality over the entire follow-up, while those aged 65 years had higher excess mortality from 3 months after diagnosis. In patients with regional cancer, the excess mortality in the oldest patients was obvious over the 2.5 first years since diagnosis and was highest in the first 12 months. For patients with distant cancer, the highest risk of death was observed during the first year since diagnosis for all ages. The excess risk in the oldest patients was visible over the entire follow-up, but highest in the first 9 months. In patients with missing stage, the excess risk of deaths was highest in the first three months regardless of age at diagnosis. The excess mortality in older patients persisted over the entire follow-up.

**Figure 2.**
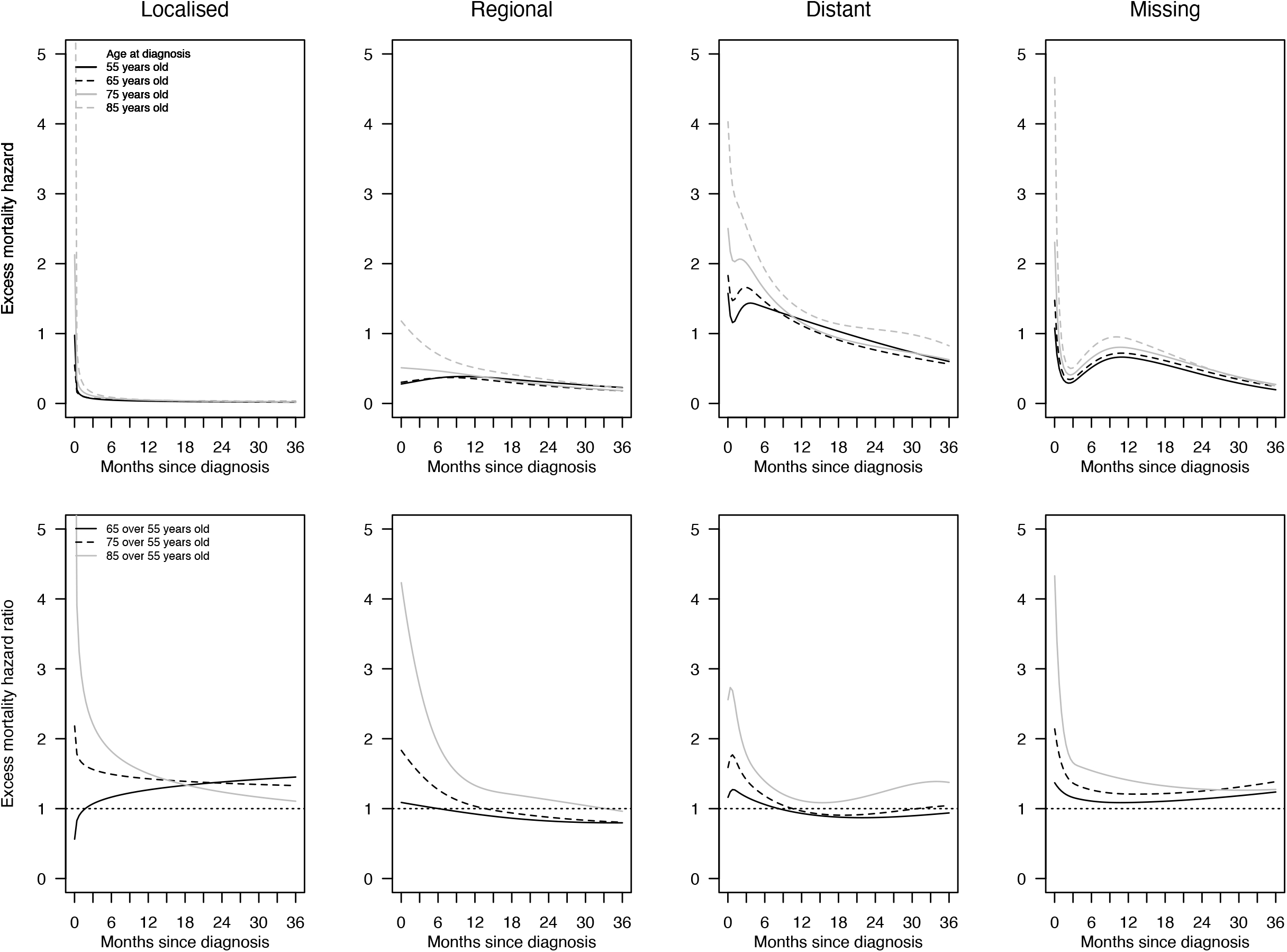
Excess mortality hazard for patients aged 55, 65, 75 and 85 years old (top panel), and excess mortality hazard ratio for patients aged 65, 75 and 85 years compared to patients aged 55 years old during the first three years since lung cancer diagnosis by stage at diagnosis (bottom panel)

Female patients younger than 75-80 years old had better net survival than males regardless of the stage at diagnosis (Figure 3). However, the sex difference disappeared around the age of 75 in patients with regional cancer, around 80-85 years in patients with distant cancer and the difference reversed from the age of 85 in patients with a missing stage. Māori patients had consistently poorer net survival than non-Māori patients. Deprivation level was not retained in the final model in patients with a missing stage (supplementary Table 3). For other stages, net survival decreased as the deprivation level increased, but the magnitude of the difference was less marked as cancer was more advanced. The role of comorbidity in net survival varied across stages. In localised cancer and missing stage, one-year net survival decreased as comorbidity level decreased. In patients with regional cancer, patients without comorbidity had slightly better one-year net survival than other patients regardless of age at diagnosis. In patients with distant cancer, the difference in net survival across comorbidity levels was small and curves crossed at 60 and 85 years showing a slighlty better one-year net survival in patients with comorbidity aged between these two ages. Patients who were diagnosed with lung cancer following an emergency presentation had consistently and significantly poorer net survival than other patients regardless of age at diagnosis and stage at diagnosis.

**Figure 3.**
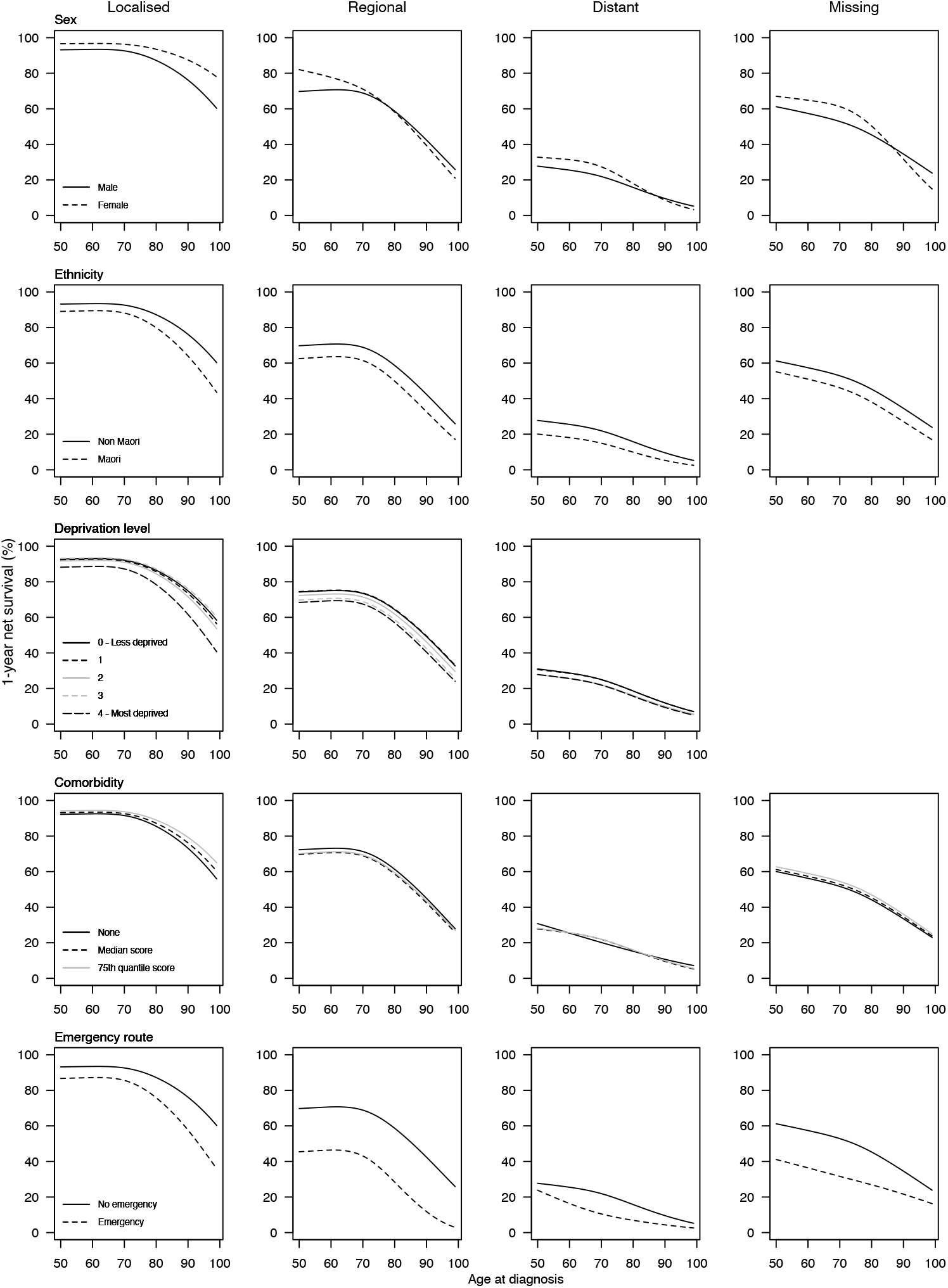
One-year net survival in patients with lung cancer aged 50-99 years by age at diagnosis, stage at diagnosis and patient-related and clinical factors

We finally tested the potential effect modification of patient-related and clinical factors on the age pattern of EMH (supplementary Table 3). No factors influenced the relationship of age on the EMH in patients with localised cancer. In patients with regional cancer, EMH ratio was higher in females at all ages, and over the entire follow-up time, but greater in the oldest women and the first three months since diagnosis (Figure 4). For distant cancer and missing stage, EMH ratio was higher in females than in males from around the age of 80, and the difference in EMH ratio between females and males was similar over time.

**Figure 4.**
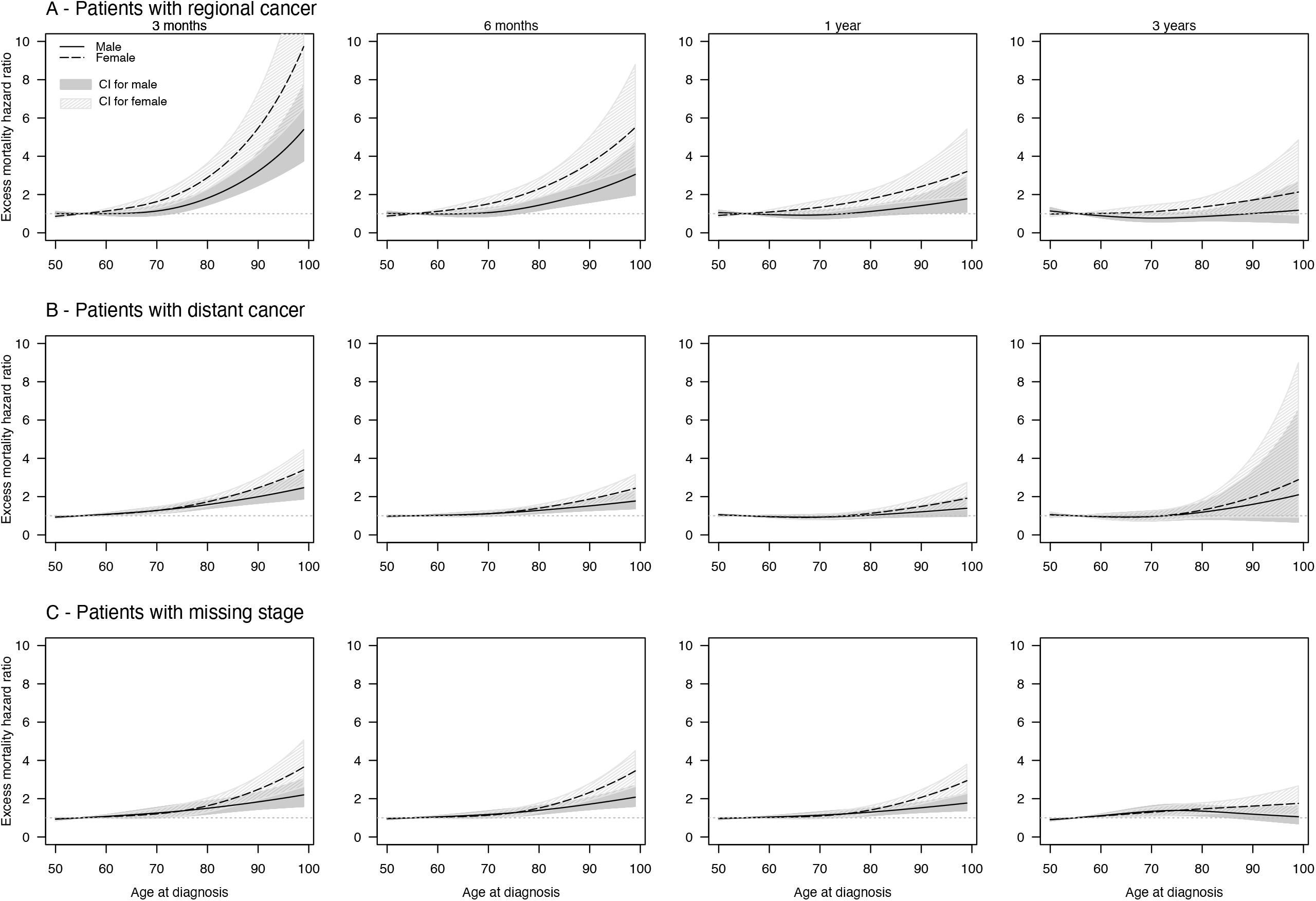
Excess mortality hazard ratios at 3, 6 months,1 and 3 years by age at diagnosis based on sex in patients with (A) regional lung cancer, (B) distant lung cancer, (C) missing stage (reference: 55 years old)

Comorbidity level interacted significantly with age at diagnosis for patients with distant cancer (Supplementary Table 3). However, the pattern of EMH ratio did not differ much across comorbidity levels (Figure 5A). An emergency presentation modified the effect of age at diagnosis on EMH in patients with distant cancer (Figure 5B) and those with a missing stage (Figure 5C). For distant cancers, patients diagnosed after an emergency presentation aged 60-80 had higher EMH ratio than those who were not, but the difference between the two groups was small. A different pattern was observed for patients with a missing stage. Patients with an emergency presentation had similar EMH ratio than other patients up to the age of 75. Then, the EMH ratio increased in patients without emergency presentation only in the first year since diagnosis before being close to one three years after diagnosis.

**Figure 5.**
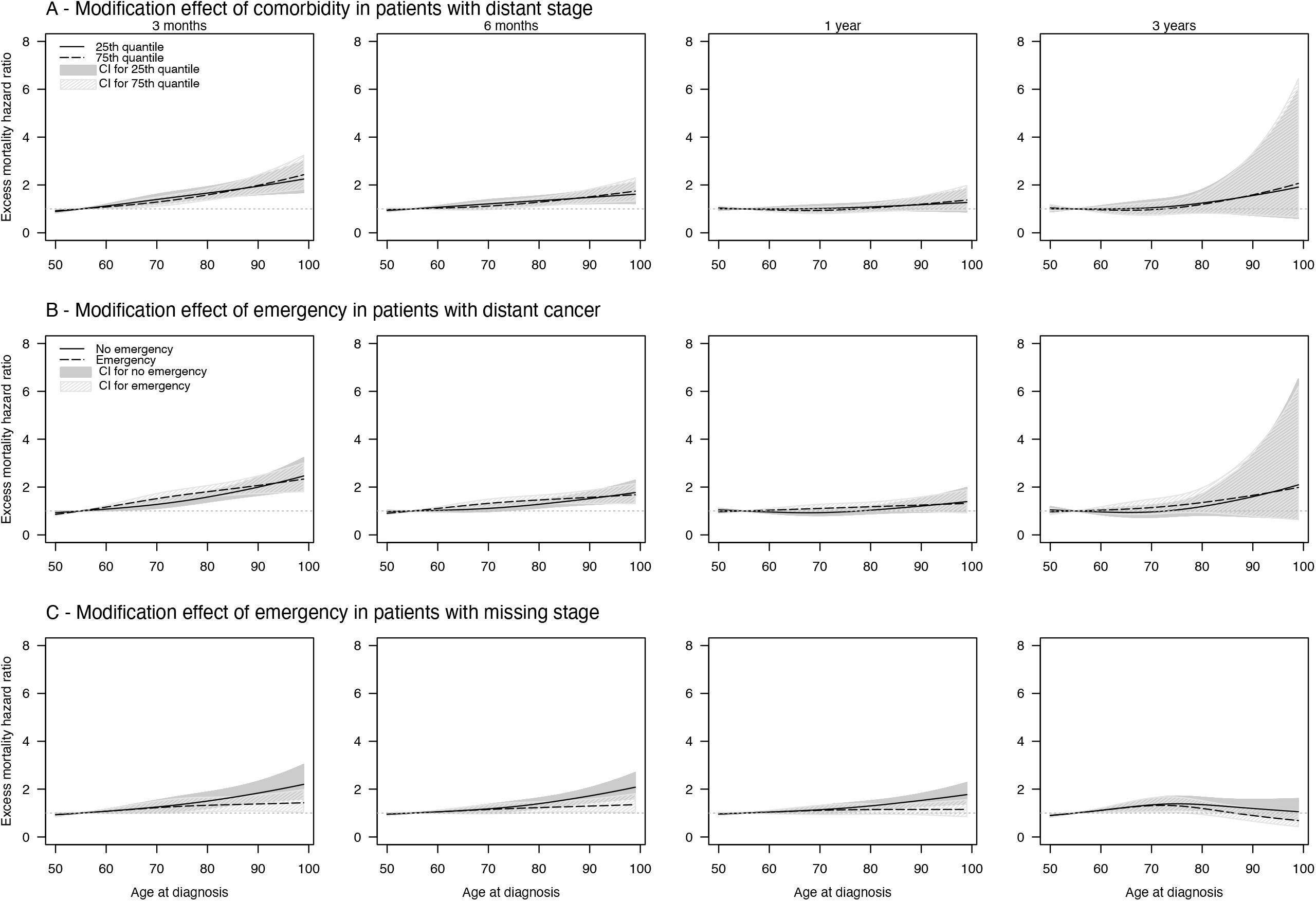
Excess mortality hazard ratios at 3, 6 months,1 and 3 years by age at diagnosis based on sex in patients with (A) regional lung cancer, (B) distant lung cancer, (C) missing stage (reference: 55 years old)

## Discussion

We described, for the first time, the role of patient-related and clinical characteristics on age pattern in net survival and excess mortality hazard in patients with lung cancer aged 50 or over. Among patient factors, female patients had greater age disparities in net survival than males. Stage at diagnosis was the main clinical factor that impacted age disparities, especially in the first year after cancer diagnosis, with bigger age disparities in non-advanced cancer. Ethnicity, deprivation level, comorbidity and an emergency presentation all explained differences in net survival, but they were not drivers of age disparities in lung cancer survival in New Zealand.

The observation of greater age disparities in lung cancer survival in females than in males is congruent with previous literature. For instance, in the study by Dickman et al., females aged 45-59 years old had better one-year relative survival than males of the same age; however, females aged 75 years or older had lower one-year relative survival than males^15^. Even if some evidence suggests a positive effect of sex hormones on survival in females^16^, the implication of sex hormones is still not clear^17^ and deserves further investigation.

Our study confirmed poorer survival in Māori patients with lung cancer and the negative role of deprivation level on lung cancer survival. However, ethnicity and deprivation level did not influence the age difference in lung cancer survival. Previous studies in the U.S and the UK suggested that race/ethnicity could have a role in age disparities, but results were inconsistent ^18,19^, probably because of differences between health-care systems. We are aware of a sole previous study on the role of socioeconomic status in age disparities in lung cancer survival, but results were not clear^20^.

Comorbidity is highly prevalent in patients with lung cancer, especially in older patients that are also more likely to have multiple comorbidities than younger patients^3^. While the interaction between comorbidity and age at diagnosis contributed significantly to the model, the actual effect of comorbidity had on age disparities was limited. A previous study presenting lung cancer survival data by age group and comorbidity level suggested greater age disparities in patients without comorbidities than those with severe comorbidities^21^. It is possible that some aspects of comorbidity were not captured by the C3 Index used to assess comorbidity level in our study. Indeed, the C3 index was created using hospitalisation data, excluding comorbidities that were not recorded during hospitalisation events, and those that did not require hospitalisation^11^. However, when compared to an index constructed using pharmaceutical data in cancer patients in New Zealand (not including lung cancer), the C3 Index performed similarly^22^. Comorbidity alone is not enough to assess vulnerabilities in older patients with cancer, and comprehensive geriatric assessments may be useful in capturing a more nuanced view of health, fitness and physiological aging^23^.

As expected and consistently with previous studies^24^, stage at diagnosis influenced age disparities in lung cancer survival. We observed the difference in survival between younger and older patients was highest in early stages of the disease, probably explained by age disparities in treatment receipt and outcomes. In later stage, the high lethality of the disease may explain the smaller difference in survival disparities observed between middle-aged and older patients. This observation with that about comorbidity above suggest that age disparities in lung cancer survival increased as cancer was amenable to treatment. The association between disparities in survival and amenability to treatment was previously described about racial disparities in cancer survival ^25^. Stage at diagnosis guides cancer treatment strategy. Surgery is recommended in fit older patients with early lung cancer^26^. For patients deemed unfit or unwilling to undergo surgery, radiotherapy is a valid option. Chemoradiation is recommended in more advanced cancers, even in older patients^26^. Yet, older patients are less likely to receive surgery, chemotherapy or radiation^6,7,27^. Some evidence suggested, however, that some of them, with appropriate stratification, may benefit from treatment^28,29^. Because of the under-representation of older patients in clinical trials^5^, clinicians lack evidence about the benefit-risk balance in older patients of potential treatment strategy and may be reluctant to offer curative treatment to their older patients. However, accumulating evidence show that comprehensive geriatric assessment (CGA) helps to identify patients who will benefit the most from treatment^30,31^. Emergency presentation, another indicator of cancer diagnosis timeliness, influenced age pattern in lung cancer survival in patients to a lesser extent. We are not aware of previous studies investigating the role of emergency presentation on age disparities in lung cancer survival. Early detection of lung cancer is likely to improve survival prospects for all patients including older ones, but this is unlikely to reduce age disparities in survival if no improvement in treatment is made for older patients.

Unfortunately, we were not able to investigate the role of treatment in this study. Because age disparities in cancer care is likely to be the main driver of age disparities in survival, it is more than urgent to improve the representation of older adults in clinical trials. Some ongoing clinical trials focus specifically on adults aged over 70 (e.g. https://clinicaltrials.gov/ct2/show/NCT03977194). Observational studies may be a good alternative to study the effect of treatment in age disparities. However, many are at high risk of bias, especially immortal time bias^24^. While waiting for clinical trials data, observational studies using appropriate methods to handle immortal time and selection biases are warranted to study the effect of treatment on age disparities in lung cancer survival^32^.

Our study has limitations. Net survival estimated in the relative survival setting relies on the accuracy of estimates of the expected mortality in general population. One assumption is that a patient with cancer would have had the same mortality risk as the general population if they had not developed a cancer. However, in the case of lung cancer, patients are more likely to smoke and have higher risk of mortality than the general population. Studies showed that lung cancer mortality hazard was overestimated, and consequently, lung cancer net survival was underestimated when not using smoking-specific life-tables^33,34^. In New Zealand, data on smoking status is collected every 7 years during population census. We did not update smoking specific life-tables for our study period because of lack of time and resources.

Another limitation comes from the fact that many lung cancers are clinically diagnosed, and the NZ PBCR receives most of its information from laboratories. This may impact especially older patients for whom clinicians may be less inclined to offer diagnostic work-up. However, the underreporting of lung cancer cases in the NZ PBCR has been estimated to be around 1-4%^35,36^.

International comparison of lung cancer survival has revealed great differences even within each stage at diagnosis^1^, especially in older patients, suggesting difference in lung cancer management across countries. Further studies in other countries are then warranted to confirm our findings or identify other factors influencing age patterns in lung cancer survival.

## Conclusion

The present population-based study confirms the importance of stage at diagnosis in age disparities in lung cancer survival, that is probably explained by age difference in lung cancer management. Our findings reinforce the call for a better representation of older adults in clinical trials and a wider use of geriatric assessment to identify patients who will benefit treatment the most to allow clinicians to make informed decisions about the best treatment strategy in this rapidly growing population^37,38^.

## Data Availability

The data that support the findings of this study are available from New Zealand Ministry of Health. Restrictions may apply to the availability of these data outside New Zealand. See https://www.health.govt.nz/nz-health-statistics/access-and-use/data-request-form.

## Acknowledgment

We thank New Zealand cancer registry staff for their help. We also thank Kendra Telfer for her immensurable help in understanding NZ datasets and Dr Nicholas West for answering our questions about cancer pathology.

## Financial disclosure

Sophie Pilleron has received funding from the European Union’s Horizon 2020 research and innovation programme under the Marie Sklodowska-Curie grant agreement No 842817.

## Declaration of competing interest

None

## Notes

### Competing Interest Statement

The authors have declared no competing interest.

### Funding Statement

This work was supported by the European Union Horizon 2020 research and innovation programme under the Marie Skłodowska-Curie grant agreement No 842817 to Sophie Pilleron.

### Author Declarations

The University of Otago Human Ethics Committee (Health) approved the study (Ethics Committee reference number HD19/048).

