## Supplementary Material for "Age disparities in lung cancer survival in New Zealand: the role of patient and clinical factors"

**Table of content**

Supplementary Table 1. Characteristics of patients with lung cancer by age group and stage at diagnosis

Supplementary Table 2. One-year and three-year net survival for patients aged 55, 65, 75, and 85 years old at lung cancer diagnosis by stage at diagnosis

Supplementary Table 3. Final excess hazard regression models by stage at diagnosis

*Model-building strategy*

We selected the best-fitting model for each stage at diagnosis. First, we selected the functional form of the baseline hazard on the most complex model yielding minimal Akaike Information Criterion (AIC). The functional form was chosen among the exponential of a restricted cubic spline and the exponential of B-spline of degree 2 or 3. For each option, we tested the number of knots (one or two) located at the median or at the tertile of the distribution of survival times in patients who died. We then selected our final models using the model building strategy of Wynant and Abrahamowicz ^1^, adapted for relative survival setting by Maringe et al.^2^ We forced the lifetable variables (*i.e.* sex, age, ethnicity, year at diagnosis) into the models as recommended ^3^. We tested the non-linear effect of age at diagnosis, year of diagnosis and comorbidity score by constructing and modelling the effects of restricted cubic splines with a knot located at the median of their distributions and boundary knots at the 10^th^ and 90^th^ percentiles of their distributions. For all covariates, we tested for time-dependent effects and their interaction with age at diagnosis using the Likelihood Ratio Test. Because of the relatively low number of events in patients with localised cancer (n=548), we aimed to keep the number of estimated parameters at its minimum. We, therefore, chose a Weibull distribution to represent the baseline hazard. We allowed only age to be non-linear, and we tested for time-dependent effects for age, sex, comorbidity and emergency presentation, and interactions of age with deprivation index, sex, ethnicity, comorbidity and emergency presentation.

**Supplementary Table 1.** Characteristics of patients with lung cancer by age group and stage at diagnosis

| **Stage at diagnosis** | **Localised** | | | | **Regional** | | | |
| --- | --- | --- | --- | --- | --- | --- | --- | --- |
| **Age category (years)** | **50-64** | **65-74** | **75-84** | **85-99** | **50-64** | **65-74** | **75-84** | **85-99** |
| Cases | 459 | 596 | 288 | 17 | 935 | 1089 | 656 | 109 |
| Person.year | 2693 | 3175 | 1279 | 51 | 2658 | 2594 | 1208 | 78 |
| Median follow-up time (years - [IQR]) | 5.50  [3.26, 8.83] | 5.11  [3.20, 7.53] | 4.02  [2.53, 6.11] | 2.27  [0.00, 5.03] | 1.92  [0.66, 4.14] | 1.42  [0.50, 3.55] | 0.86  [0.24, 2.58] | 0.27  [0.07, 0.73] |
| Deaths (%) | 146 (31.8) | 239 (40.1) | 148 (51.4) | 15 (88.2) | 694 (74.2) | 867 (79.6) | 562 (85.7) | 106 (97.2) |
| **Patient-related factors** |  |  |  |  |  |  |  |  |
| Male (%) | 194 (42.3) | 295 (49.5) | 148 (51.4) | 10 (58.8) | 435 (46.5) | 586 (53.8) | 382 (58.2) | 62 (56.9) |
| Ethnicity (%) |  |  |  |  |  |  |  |  |
| Māori | 89 (19.4) | 70 (11.7) | 30 (10.4) | 0 (0.0) | 304 (32.5) | 207 (19.0) | 68 (10.4) | 8 (7.3) |
| Non-Māori | 370 (80.6) | 526 (88.3) | 258 (89.6) | 17 (100.0) | 631 (67.5) | 882 (81.0) | 588 (89.6) | 101 (92.7) |
| Deprivation Index Quintiles (%) |  |  |  |  |  |  |  |  |
| 1 – Less deprived | 69 (15.0) | 85 (14.3) | 47 (16.3) | 2 (11.8) | 116 (12.4) | 149 (13.7) | 89 (13.6) | 10 (9.2) |
| 2 | 56 (12.2) | 93 (15.6) | 42 (14.6) | 2 (11.8) | 140 (15.0) | 157 (14.4) | 103 (15.7) | 17 (15.6) |
| 3 | 103 (22.4) | 125 (21.0) | 57 (19.8) | 4 (23.5) | 164 (17.5) | 244 (22.4) | 132 (20.1) | 20 (18.3) |
| 4 | 106 (23.1) | 143 (24.0) | 86 (29.9) | 4 (23.5) | 218 (23.3) | 237 (21.8) | 164 (25.0) | 33 (30.3) |
| 5 - More deprived | 125 (27.2) | 150 (25.2) | 56 (19.4) | 5 (29.4) | 297 (31.8) | 302 (27.7) | 168 (25.6) | 29 (26.6) |
| **Clinical factors** |  |  |  |  |  |  |  |  |
| Comorbidity score >0 (%) | 296 (64.5) | 433 (72.7) | 207 (71.9) | 14 (82.4) | 680 (72.7) | 819 (75.2) | 519 (79.1) | 72 (66.1) |
| Comorbidity score (median [IQR]) | 0.98  [0.00, 1.88] | 1.09  [0.00, 2.67] | 1.09  [0.00, 2.46] | 1.35  [0.52, 1.84] | 1.09  [0.00, 1.92] | 1.13  [0.11, 2.53] | 1.23  [0.52, 2.55] | 1.10  [0.00, 2.93] |
| Emergency presentation (%) | 57 (12.4) | 57 (9.6) | 26 (9.0) | 5 (29.4) | 262 (28.0) | 287 (26.4) | 205 (31.2) | 57 (52.3) |

**Supplementary Table 1 (continued)**. Characteristics of patients with lung cancer by age group and stage at diagnosis

| **Stage at diagnosis** | **Distant** | | | | **Missing** | | | |
| --- | --- | --- | --- | --- | --- | --- | --- | --- |
| **Age category (years)** | **50-64** | **65-74** | **75-84** | **85-99** | **50-64** | **65-74** | **75-84** | **85-99** |
| Cases | 3039 | 3432 | 2649 | 874 | 1629 | 2618 | 2905 | 1192 |
| Person.year | 2485 | 2256 | 1202 | 285 | 3042 | 4161 | 3541 | 1058 |
| Median follow-up time (years - [IQR]) | 0.36  [0.14, 0.80] | 0.26  [0.10, 0.65] | 0.17  [0.07, 0.39] | 0.13  [0.06,0.35] | 0.98  [0.40, 2.10] | 0.84  [0.29, 1.96] | 0.61  [0.15, 1.48] | 0.31  [0.06,0.94] |
| Deaths (%) | 2930 (96.4) | 3343 (97.4) | 2609 (98.5) | 873 (99.9) | 1423 (87.4) | 2386 (91.1) | 2783 (95.8) | 1169 (98.1) |
| **Patient-related factors** |  |  |  |  |  |  |  |  |
| Male (%) | 1513 (49.8) | 1841 (53.6) | 1475 (55.7) | 482 (55.1) | 783 (48.1) | 1421 (54.3) | 1627 (56.0) | 633 (53.1) |
| Ethnicity (%) |  |  |  |  |  |  |  |  |
| Māori | 913 (30.0) | 650 (18.9) | 258 (9.7) | 34 (3.9) | 615 (37.8) | 637 (24.3) | 375 (12.9) | 51 (4.3) |
| Non-Māori | 2126 (70.0) | 2782 (81.1) | 2391 (90.3) | 840 (96.1) | 1014 (62.2) | 1981 (75.7) | 2530 (87.1) | 1141 (95.7) |
| Deprivation Index Quintiles (%) |  |  |  |  |  |  |  |  |
| 1 – Less deprived | 345 (11.4) | 430 (12.5) | 324 (12.2) | 125 (14.3) | 150 (9.2) | 237 (9.1) | 319 (11.0) | 134 (11.2) |
| 2 | 428 (14.1) | 485 (14.1) | 386 (14.6) | 161 (18.4) | 208 (12.8) | 336 (12.8) | 421 (14.5) | 216 (18.1) |
| 3 | 541 (17.8) | 668 (19.5) | 571 (21.6) | 200 (22.9) | 290 (17.8) | 484 (18.5) | 600 (20.7) | 275 (23.1) |
| 4 | 730 (24.0) | 876 (25.5) | 723 (27.3) | 218 (24.9) | 386 (23.7) | 707 (27.0) | 785 (27.0) | 310 (26.0) |
| 5 - More deprived | 995 (32.7) | 973 (28.4) | 645 (24.3) | 170 (19.5) | 595 (36.5) | 854 (32.6) | 780 (26.9) | 257 (21.6) |
| **Clinical factors** |  |  |  |  |  |  |  |  |
| Comorbidity score >0 (%) | 1929 (63.5) | 2382 (69.4) | 1958 (73.9) | 634 (72.5) | 1072 (65.8) | 1921 (73.4) | 2267 (78.0) | 911 (76.4) |
| Comorbidity score (median [IQR]) | 0.75  [0.00, 1.70] | 1.08  [0.00, 2.12] | 1.09  [0.00, 2.44] | 1.09  [0.00,2.24] | 1.09  [0.00, 2.06] | 1.09  [0.00, 2.47] | 1.39  [0.61, 2.92] | 1.38  [0.51,3.09] |
| Emergency presentation (%) | 1622 (53.4) | 1771 (51.6) | 1482 (55.9) | 522 (59.7) | 511 (31.4) | 797 (30.4) | 1005 (34.6) | 462 (38.8) |

**Supplementary Table 2.** One-year and three-year net survival for patients aged 55, 65, 75, and 85 years old at lung cancer diagnosis by stage at diagnosis

|  | Localised | | Regional | | Distant | | Missing | |
| --- | --- | --- | --- | --- | --- | --- | --- | --- |
|  | 1-year NS | 3-year NS | 1-year NS | 3-year NS | 1-year NS | 3-year NS | 1-year NS | 3-year NS |
| 55 years old | 93.3 (89.1 - 95.9) | 88.8 (83.0 - 92.7) | 70.3 (64.9 – 75.0) | 38.3 (31.4 - 45.1) | 26.7 (22.8 - 30.7) | 4.6 (2.8 - 6.9) | 59.3 (55.2 - 63.2) | 25.5 (21.5 - 29.8) |
| 65 years old | 93.2 (90.4 - 95.3) | 87.2 (82.3 - 90.8) | 70.3 (66.4 - 73.9) | 42.2 (36.9 - 47.3) | 24.0 (21.3 - 26.8) | 5.0 (3.5 - 6.9) | 55.3 (52.8 - 57.7) | 21.3 (19.0 - 23.6) |
| 75 years old | 89.3 (85.2 - 92.3) | 83.4 (77.5 - 87.9) | 63.1 (58.6 - 67.2) | 36.5 (31.4 - 41.6) | 19.0 (16.6 - 21.6) | 3.6 (2.4 - 5.1) | 49.6 (47.0 - 52.1) | 17.3 (15.2 - 19.5) |
| 85 years old | 78.7 (67.4 - 86.5) | 73.9 (60.8 - 83.2) | 47.2 (39.3 - 54.6) | 23.6 (16.5 - 31.4) | 12.5 (9.9 - 15.4) | 1.5 (0.7 - 2.8) | 40.2 (37.1 - 43.2) | 13.3 (11.1 - 15.7) |

**Supplementary Table 3.** Final excess hazard regression models by stage at diagnosis

| Stage at diagnosis | Localised | | | | Regional | | | | Distant | | | | Missing | | | |
| --- | --- | --- | --- | --- | --- | --- | --- | --- | --- | --- | --- | --- | --- | --- | --- | --- |
| Baseline Hazard functional form | Weibull distribution | | | | Exponential of restricted spline with two knots at tertile of survival time in patients who died | | | | Exponential of cubic B-spline with two knots at tertile of survival time in patients who died | | | | Exponential of cubic B-spline with two knots at tertile of survival time in patients who died | | | |
|  | Main effect | Non linearity^#^ | Time-dependent effect | Modification effect by age | Main effect | Non linearity^#^ | Time-dependent effect | Modification effect by age | Main effect | Non linearity^#^ | Time-dependent effect | Modification effect by age | Main effect | Non linearity^#^ | Time-dependent effect | Modification effect by age |
| Age  (continuous - forced) | X  p<0.001 | X  p<0.001 | X  p<0.001 | N/A | X  p<0.001 | X  p=0.003 | X  p<0.001 | N/A | X  p<0.001 | X  p=0.003 | X  p<0.001 | N/A | X  p<0.001 | X  p<0.001 | X  p<0.001 | N/A |
| Year  (continuous -forced) | X  p<0.001 | Not tested | Not tested | Not tested | X  p<0.001 | X  p<0.001 |  |  | X  p<0.001 | X  p<0.001 | X  p=0.002 |  | X  p<0.001 | X  p<0.001 | X  p<0.001 |  |
| Sex  (dichotomy-forced) | X  P<0.001 | N/A | X  p=0.002 |  | X  p<0.001 | N/A |  | X  p<0.001 | X  p<0.001 | N/A | X  p<0.001 | X  p=0.015 | X  p<0.001 | N/A | X  p=0.020 | X  p<0.001 |
| Ethnicity  (dichotomy-forced) | X  p=0.003 | N/A | Not tested |  | X  p<0.001 | N/A |  |  | X  p<0.001 | N/A |  |  | X  p<0.001 | N/A | X  p=0.048 |  |
| Deprivation (categorical) | X  p=0.008 | N/A | Not tested |  | X  p=0.001 | N/A |  |  | X  p<0.001 | N/A | X  p<0.001 |  |  |  |  |  |
| Comorbidity (continuous) | X  p<0.001 | Not tested | Did not converge |  | X  p<0.001 | X  p=0.018 | X  p<0.001 |  | X  p<0.001 | X  p<0.001 | X  p<0.001 | X  p=0.045 | X  p<0.001 |  | X  p<0.001 |  |
| Emergency (dichotomy) | X  p=0.002 | N/A | X  p=0.006 |  | X  p<0.001 | N/A | X  p<0.001 |  | X  p<0.001 | N/A | X  p<0.001 | X  p=0.011 | X  p<0.001 | N/A | X  p<0.001 | X  p=0.004 |
| Observations | 1360 | | | | 2789 | | | | 9994 | | | | 8344 | | | |
| Events | 548 | | | | 2229 | | | | 9755 | | | | 8252 | | | |
| # events/ # parameters ratio | 30 | | | | 66 | | | | 112 | | | | 142 | | | |

X: term retained in the final models; N/A: Not applicable

^#^ Non-linearity: Restricted cubic spline with one knot located at the median and boundaries knots located at 10th and 90th percentile

p-values obtained using Log Likelihood ratio test
